# Postoperative mortality analysis of national Japanese Diagnosis Procedure Combination database with a focus on regional comparisons and changes over time

**DOI:** 10.1101/2022.09.09.22279570

**Authors:** Susumu Kunisawa

## Abstract

**PURPOSE:** This study aimed to investigate postoperative mortality from all surgeries at the prefecture level using nationwide database, Japanese Diagnosis Procedure Combination (DPC) database, and to show those with visible changes over time or variations among areas.

**METHODS:** Data were provided in accordance with the guidelines and application as indicated on the Ministry of Health, Labor and Welfare (MHLW), Japan. The number of cases and in-hospital mortality were calculated for each representative surgery for each hospitalization by fiscal year of discharge from 2011 to 2018 and by prefecture. Values of ≥10 in each aggregated data cell are presented.

**RESULTS:** The aggregated result data contain 474,154 records, with about 2000 different surgical codes. Only in the 16,890 data, more than 10 mortalities were recorded, which can be used in the mortality analysis. In the analyses of artificial head insertion, cerebral aneurysm neck clipping, coronary artery and aortic bypass grafting, and tracheotomy, regional differences and a declining trend were observed in some categories.

**CONCLUSION:** In addition to considering categories that can be used in the analysis, careful consideration must be given to the inclusion of background context such as quality of care.

## Introduction

Improving the quality of care is one of the items of greatest concern in the medical community, and reducing postoperative complications deserves attention. Various case registries have been conducted, verified, and reviewed, mainly at academic conferences [1,2]. Currently, huge case data have been accumulated in the Japanese surgical field, mainly through the National Clinical Database (NCD), and these data are being used for validation and feedback to facilities. However, what can be obtained from such a case registry is limited to the data registered. Although some surgical procedures have fairly high registration rates [3], registration must be a proactive approach; thus, registration omissions can still occur. In addition, the NCD (https://www.ncd.or.jp/press/), for example, do not necessarily include data from all areas of surgery, although they are being used in major surgeries such as gastroenterology, cardiovascular surgery, respiratory surgery, and pediatric surgery [4–6]. This can be one of the limitations based on any case registries. In Japan, along with the evolution of information technology, the Diagnosis Procedure Combination (DPC) data through the DPC per-diem payment system (DPC/PDPS), which started in 2003, was created in parallel with the accumulation and utilization of DPC data. DPC data should be prepared as a kind of discharge summary called “Form 1” in addition to recording at the voucher level the medical treatment activities performed during treatment, despite being a comprehensive payment system. Initially, these data were defined for the DPC/PDPS system, but as the requirements for creating such a database have been extended to medical institutions that have not adopted DPC/PDPS, the DPC database has now been created and submitted to the Ministry of Health, Labor and Welfare (MHLW) by many hospitals (5315 hospitals in 2020; https://www.mhlw.go.jp/stf/shingi2/0000196043_00005.html).

The DPC database, which holds data on the vast majority of hospitalizations related to insured care, has been applied on practical management analyses and research for clinical analysis from various perspectives. The DPC database has been very useful for research across organizations because it contains government-defined data. There have been many intra-hospital analyses and cross-sectional or inter-hospital analyses within multiple organizations or groups [7–11]. Moreover, the use of data collected by the MHLW has become possible in 2017. Previously, a study successfully calculated process indicators using this database [12], which clearly showed regional differences and changes over time in Japan.

Postoperative mortality can be considered one of the quality indicators. Although mortality has various meanings for backgrounds such as not only the technical quality but also the difference in case-mix and indication strategies, the mortality rates in Japan often are lower than those in other countries [13,14]. However, such results are obtained from specific hospitals or registry data analysis. Thus, the present study was designed to explore regional differences and changes over time in postoperative mortality for any surgeries as an outcome measure within the quality of care using a more comprehensive nationwide data from Japan. Rather than examining details such as the causes of regional differences in specific surgeries, this study fundamentally calculated and provided these indicators, thereby providing a very important foundation for ideas that can be gleaned from them and for more detailed and specific studies in the future. Through this analysis, this paper presents some caveats for the use of data for cross-disciplinary indicators and reports several items that should be cared for by future investigators.

## Methods

Data were provided in accordance with the guidelines and application as indicated on the MHLW. The number of cases and in-hospital mortality were calculated for each representative surgery for each hospitalization by fiscal year of discharge from 2011 to 2018 and by prefecture, and data were provided as an aggregated result from the MHLW. The DPC data “Form 1” allows recording of a maximum of five surgeries, and the primary surgery or the one with the highest surgical fee should be recorded as the first surgery. Surgical information recorded included the surgical name and surgical code (K-code). Since the surgical name causes some notational inconsistencies, data were tabulated by surgical code, and the resulting data were then combined with the standard master surgery names for those codes provided. Surgery names are in Japanese, the master data are available at Various Information of Medical Fee (http://shinryohoshu.mhlw.go.jp/shinryohoshu/), and the referable translation is available at the Japan Surgical Society – Web Glossary of Surgical Terms (http://yougosyu.jssoc.or.jp/). Data that can be “obtained” (not “disclosed to this report”) as an aggregate was limited by the rules of MHLW to a value of ≥10 in each cell, and if the number was less, it was removed and indicated “-.” In addition, the number was removed and indicated “*-” if the number of cases minus the number of in-hospital mortality was <10. In this study, postoperative in-hospital mortality was defined and calculated as the number of in-hospital mortality divided by the number of postoperative case discharges. Therefore, patients who were discharged alive once and died during such as readmission were not included in the mortality data.

The number of DPC-participating hospitals is increasing annually. As mentioned, submission of DPC data has been expanded to non-DPC hospitals in recent years, and the later the period, the larger the number of hospitals. Consequently, more cases are being covered by this database, so the case volume could increase.

All surgery data obtained in this study will be presented as supplements, and an arbitrary selection of some of the data will be presented in the Results. This selection was made arbitrarily by the author, but the inclusion criteria were as follows: approximately those with relatively high mortality rates, those with visible changes over time or variations among areas, those that require interpretation, or those that require caution in future data analysis.

This study was approved by Kyoto University Graduate School and Faculty of Medicine, Ethics Committee. The data presented in this study were independently requested and analyzed by the author and are different from the statistics, etc., prepared and published by the MHLW Japan.

## Results

All data obtained in this study are presented as supplements, except for the unknown operation code data of 24 records. The dataset contains 474,154 records, and for each year, the dataset contained 1986–2496 different codes. Those with zero cases were not included in the dataset, and the 221,866 records had more than 10 cases. Only the 16890 records had >10 mortalities, and 186 records were masked along the methods. With just 56 surgery codes, the mortality rates for all prefectures during the fiscal year were comparable, and with just 1063 surgery codes, the mortality rates during a fiscal year were comparable among prefectures.

### Artificial head insertion for the shoulder or hip (K0811)

Table 1 shows the results for cases of artificial head insertion for the shoulder or hip in orthopedic surgery. Figure 1 graphically depicts the postoperative in-hospital mortality. Surgeries coded with K0811 are artificial head insertions for the shoulder or hip; thus, the cases are mixed. Moreover, the rate of postoperative in-hospital mortality was approximately 1%–3% and appeared to vary somewhat regionally. However, there are many areas where numerical aggregation is impossible because of the small number of postoperative in-hospital mortalities in each area.

**Table 1.** Artificial head insertion (K0811), case number, and outcomes of postoperative in-hospital mortality

| Fiscal year | 2011 |  |  | 2012 |  |  | 2013 |  |  | 2014 |  |  | 2015 |  |  | 2016 |  |  | 2017 |  |  | 2018 |  |  |
| --- | --- | --- | --- | --- | --- | --- | --- | --- | --- | --- | --- | --- | --- | --- | --- | --- | --- | --- | --- | --- | --- | --- | --- | --- |
| Prefecture | Case | Outcome |  | Case | Outcome |  | Case | Outcome |  | Case | Outcome |  | Case | Outcome |  | Case | Outcome |  | Case | Outcome |  | Case | Outcome |  |
| 01 | 1221 | 20 | 1.6% | 1358 | 26 | 1.9% | 1436 | 27 | 1.9% | 1692 | 21 | 1.2% | 1978 | 42 | 2.1% | 2119 | 38 | 1.8% | 2254 | 30 | 1.3% | 2406 | 43 | 1.8% |
| 02 | 286 | - | - | 321 | - | - | 286 | - | - | 342 | - | - | 378 | - | - | 432 | - | - | 477 | 11 | 2.3% | 500 | - | - |
| 03 | 319 | - | - | 334 | - | - | 329 | - | - | 368 | 14 | 3.8% | 399 | - | - | 364 | - | - | 453 | - | - | 528 | - | - |
| 04 | 555 | - | - | 536 | - | - | 486 | - | - | 619 | - | - | 613 | - | - | 744 | - | - | 741 | 12 | 1.6% | 816 | - | - |
| 05 | 306 | - | - | 265 | - | - | 290 | - | - | 312 | - | - | 357 | - | - | 352 | - | - | 385 | - | - | 371 | - | - |
| 06 | 390 | - | - | 375 | - | - | 363 | - | - | 379 | - | - | 405 | 10 | 2.5% | 378 | - | - | 452 | - | - | 435 | - | - |
| 07 | 551 | 10 | 1.8% | 550 | 12 | 2.2% | 556 | 11 | 2.0% | 626 | 12 | 1.9% | 676 | 12 | 1.8% | 715 | 10 | 1.4% | 808 | 18 | 2.2% | 849 | 16 | 1.9% |
| 08 | 775 | 18 | 2.3% | 796 | 24 | 3.0% | 683 | 15 | 2.2% | 895 | 24 | 2.7% | 928 | 15 | 1.6% | 1032 | 20 | 1.9% | 1204 | 26 | 2.2% | 1221 | 32 | 2.6% |
| 09 | 366 | - | - | 424 | 11 | 2.6% | 490 | 14 | 2.9% | 581 | 16 | 2.8% | 677 | 12 | 1.8% | 687 | 18 | 2.6% | 784 | 12 | 1.5% | 877 | 18 | 2.1% |
| 10 | 411 | - | - | 499 | - | - | 498 | - | - | 579 | 12 | 2.1% | 664 | - | - | 702 | - | - | 763 | - | - | 691 | - | - |
| 11 | 1420 | 27 | 1.9% | 1535 | 37 | 2.4% | 1648 | 28 | 1.7% | 1954 | 38 | 1.9% | 2289 | 37 | 1.6% | 2651 | 54 | 2.0% | 2834 | 58 | 2.0% | 3094 | 78 | 2.5% |
| 12 | 1173 | 33 | 2.8% | 1269 | 34 | 2.7% | 1321 | 24 | 1.8% | 1650 | 25 | 1.5% | 1855 | 47 | 2.5% | 2267 | 40 | 1.8% | 2334 | 44 | 1.9% | 2526 | 59 | 2.3% |
| 13 | 3709 | 58 | 1.6% | 3843 | 73 | 1.9% | 4198 | 60 | 1.4% | 4720 | 75 | 1.6% | 5049 | 83 | 1.6% | 5341 | 91 | 1.7% | 5957 | 124 | 2.1% | 6230 | 101 | 1.6% |
| 14 | 2860 | 51 | 1.8% | 3047 | 56 | 1.8% | 3358 | 43 | 1.3% | 3721 | 82 | 2.2% | 4050 | 78 | 1.9% | 4300 | 89 | 2.1% | 4547 | 79 | 1.7% | 4706 | 81 | 1.7% |
| 15 | 547 | - | - | 619 | 11 | 1.8% | 720 | 14 | 1.9% | 785 | 22 | 2.8% | 990 | 20 | 2.0% | 975 | 14 | 1.4% | 1136 | 17 | 1.5% | 1117 | 13 | 1.2% |
| 16 | 407 | - | - | 412 | - | - | 459 | - | - | 433 | - | - | 564 | - | - | 540 | - | - | 549 | 13 | 2.4% | 516 | - | - |
| 17 | 370 | 11 | 3.0% | 350 | - | - | 331 | - | - | 404 | - | - | 446 | - | - | 434 | - | - | 481 | 14 | 2.9% | 504 | 16 | 3.2% |
| 18 | 297 | - | - | 339 | - | - | 342 | - | - | 356 | 11 | 3.1% | 315 | - | - | 327 | - | - | 344 | - | - | 381 | - | - |
| 19 | 253 | 10 | 4.0% | 267 | - | - | 275 | - | - | 282 | - | - | 306 | - | - | 365 | - | - | 425 | - | - | 469 | - | - |
| 20 | 686 | 10 | 1.5% | 763 | 11 | 1.4% | 788 | 14 | 1.8% | 904 | 12 | 1.3% | 943 | 14 | 1.5% | 960 | 24 | 2.5% | 1047 | 20 | 1.9% | 1056 | 16 | 1.5% |
| 21 | 905 | 14 | 1.5% | 909 | 15 | 1.7% | 857 | 17 | 2.0% | 1050 | 24 | 2.3% | 1107 | 27 | 2.4% | 1068 | 15 | 1.4% | 1217 | 22 | 1.8% | 1235 | 32 | 2.6% |
| 22 | 1522 | 24 | 1.6% | 1509 | 29 | 1.9% | 1555 | 20 | 1.3% | 1727 | 21 | 1.2% | 1774 | 25 | 1.4% | 1891 | 21 | 1.1% | 2022 | 29 | 1.4% | 1985 | 33 | 1.7% |
| 23 | 1796 | 26 | 1.4% | 2052 | 23 | 1.1% | 2254 | 29 | 1.3% | 2424 | 46 | 1.9% | 2758 | 43 | 1.6% | 2872 | 38 | 1.3% | 3006 | 49 | 1.6% | 3024 | 49 | 1.6% |
| 24 | 693 | 12 | 1.7% | 704 | - | - | 757 | 17 | 2.2% | 823 | 19 | 2.3% | 923 | 18 | 2.0% | 975 | - | - | 1160 | 26 | 2.2% | 1123 | 27 | 2.4% |
| 25 | 462 | - | - | 453 | 10 | 2.2% | 501 | 10 | 2.0% | 526 | - | - | 598 | - | - | 654 | 19 | 2.9% | 692 | 15 | 2.2% | 670 | 16 | 2.4% |
| 26 | 790 | 19 | 2.4% | 965 | 29 | 3.0% | 978 | 27 | 2.8% | 1082 | 29 | 2.7% | 1198 | 24 | 2.0% | 1215 | 37 | 3.0% | 1364 | 32 | 2.3% | 1423 | 21 | 1.5% |
| 27 | 2986 | 70 | 2.3% | 3280 | 85 | 2.6% | 3376 | 87 | 2.6% | 3961 | 99 | 2.5% | 4426 | 83 | 1.9% | 4593 | 116 | 2.5% | 4973 | 124 | 2.5% | 5376 | 122 | 2.3% |
| 28 | 1764 | 39 | 2.2% | 1910 | 34 | 1.8% | 1991 | 31 | 1.6% | 2302 | 53 | 2.3% | 2815 | 64 | 2.3% | 2996 | 65 | 2.2% | 3119 | 66 | 2.1% | 3225 | 83 | 2.6% |
| 29 | 618 | 18 | 2.9% | 596 | 15 | 2.5% | 685 | - | - | 628 | 15 | 2.4% | 742 | 16 | 2.2% | 764 | 20 | 2.6% | 837 | 17 | 2.0% | 1031 | 17 | 1.6% |
| 30 | 489 | - | - | 545 | 15 | 2.8% | 523 | - | - | 612 | 13 | 2.1% | 610 | - | - | 668 | 11 | 1.6% | 767 | 24 | 3.1% | 734 | 19 | 2.6% |
| 31 | 234 | - | - | 305 | - | - | 266 | - | - | 295 | - | - | 403 | - | - | 382 | - | - | 370 | - | - | 418 | 11 | 2.6% |
| 32 | 240 | - | - | 313 | - | - | 293 | - | - | 349 | - | - | 371 | - | - | 393 | - | - | 465 | - | - | 408 | 10 | 2.5% |
| 33 | 681 | - | - | 805 | - | - | 796 | 20 | 2.5% | 896 | 12 | 1.3% | 1021 | 13 | 1.3% | 1064 | 19 | 1.8% | 1202 | 19 | 1.6% | 1147 | 19 | 1.7% |
| 34 | 998 | 17 | 1.7% | 1076 | 24 | 2.2% | 1112 | 20 | 1.8% | 1246 | 30 | 2.4% | 1368 | 32 | 2.3% | 1511 | 33 | 2.2% | 1606 | 29 | 1.8% | 1743 | 44 | 2.5% |
| 35 | 612 | 10 | 1.6% | 653 | 12 | 1.8% | 680 | - | - | 782 | 12 | 1.5% | 843 | 14 | 1.7% | 865 | 20 | 2.3% | 947 | 16 | 1.7% | 997 | 15 | 1.5% |
| 36 | 322 | - | - | 411 | - | - | 405 | - | - | 524 | - | - | 554 | - | - | 621 | - | - | 587 | - | - | 592 | - | - |
| 37 | 390 | - | - | 411 | - | - | 430 | - | - | 432 | - | - | 463 | - | - | 466 | - | - | 559 | - | - | 557 | - | - |
| 38 | 355 | - | - | 396 | - | - | 452 | 14 | 3.1% | 593 | 10 | 1.7% | 687 | - | - | 722 | 14 | 1.9% | 794 | - | - | 878 | 18 | 2.1% |
| 39 | 329 | - | - | 353 | - | - | 356 | - | - | 369 | - | - | 388 | - | - | 369 | - | - | 457 | 15 | 3.3% | 456 | 11 | 2.4% |
| 40 | 1991 | 24 | 1.2% | 2071 | 28 | 1.4% | 2136 | 41 | 1.9% | 2344 | 42 | 1.8% | 2537 | 33 | 1.3% | 2750 | 34 | 1.2% | 2906 | 61 | 2.1% | 2953 | 54 | 1.8% |
| 41 | 320 | - | - | 334 | - | - | 341 | - | - | 389 | - | - | 485 | - | - | 481 | - | - | 498 | - | - | 473 | - | - |
| 42 | 657 | 11 | 1.7% | 674 | - | - | 701 | - | - | 818 | 14 | 1.7% | 833 | 14 | 1.7% | 902 | 15 | 1.7% | 1002 | 15 | 1.5% | 1026 | 14 | 1.4% |
| 43 | 935 | - | - | 959 | 11 | 1.1% | 938 | 14 | 1.5% | 1031 | 15 | 1.5% | 1046 | - | - | 1192 | 22 | 1.8% | 1184 | 13 | 1.1% | 1150 | 15 | 1.3% |
| 44 | 400 | 11 | 2.8% | 521 | - | - | 523 | - | - | 551 | 14 | 2.5% | 723 | 13 | 1.8% | 693 | 15 | 2.2% | 782 | 10 | 1.3% | 755 | 14 | 1.9% |
| 45 | 392 | - | - | 396 | - | - | 413 | - | - | 561 | - | - | 596 | - | - | 627 | - | - | 690 | - | - | 655 | 10 | 1.5% |
| 46 | 519 | 14 | 2.7% | 560 | 14 | 2.5% | 488 | 15 | 3.1% | 637 | 15 | 2.4% | 735 | 20 | 2.7% | 816 | 11 | 1.3% | 903 | 10 | 1.1% | 960 | - | - |
| 47 | 624 | - | - | 630 | - | - | 683 | - | - | 692 | 10 | 1.4% | 712 | - | - | 759 | - | - | 736 | 10 | 1.4% | 743 | - | - |

- indicates <10.

**Fig. 1.**
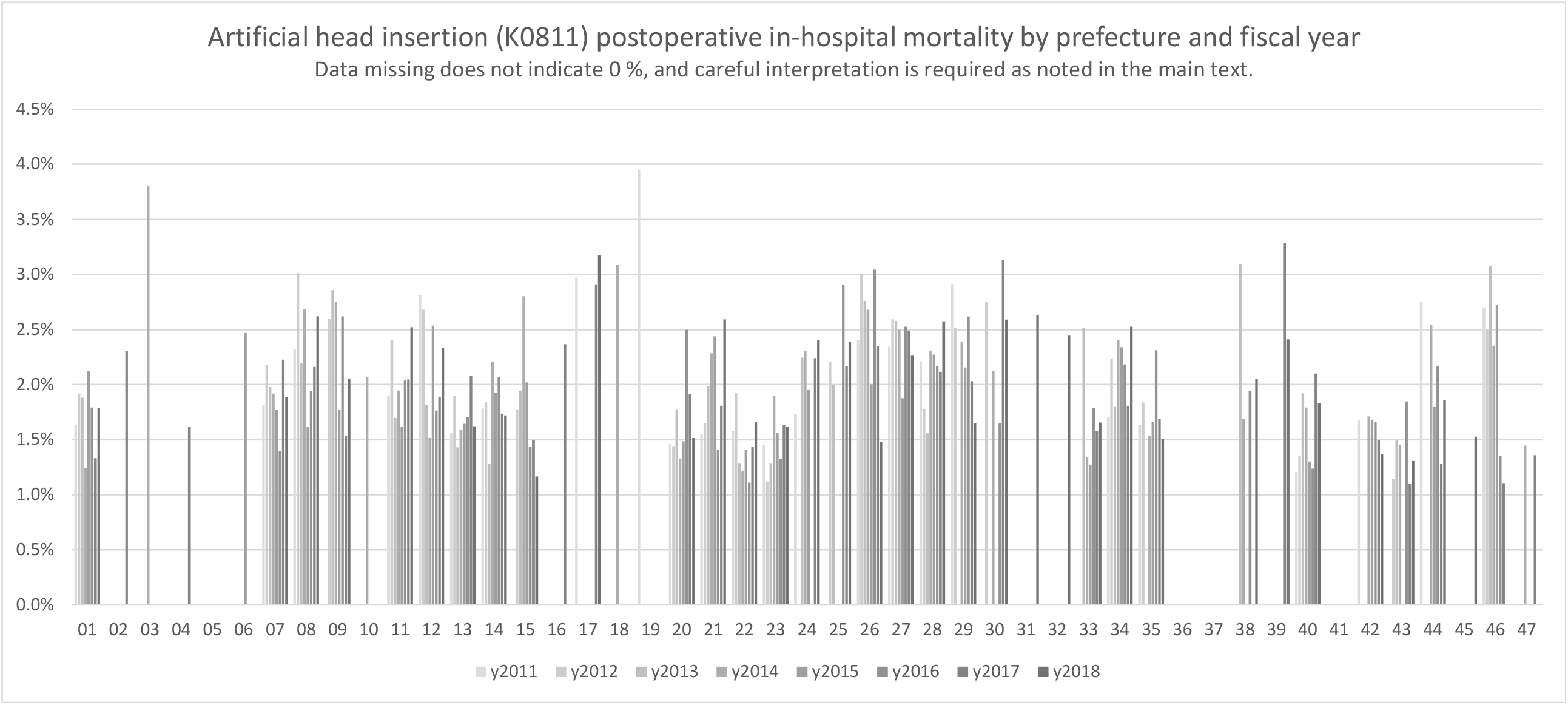
Artificial head insertion (K0811)

### Cerebral aneurysm neck clipping (1 location) (K1771)

Table 2 shows the results for cases of cerebral aneurysm neck clipping. This surgery code is used to indicate clipping of one location, and multiple location surgery is coded K1772. Figure 2 graphically depicts the postoperative in-hospital mortality. A decreasing trend over time can be seen in 14 (Miyagi Prefecture), 22 (Shizuoka Prefecture), and 23 (Aichi Prefecture).

**Table 2.** Cerebral aneurysm neck clipping (1 location) (K1771), case number, and outcomes of postoperative in-hospital mortality

| Fiscal year | 2011 |  |  | 2012 |  |  | 2013 |  |  | 2014 |  |  | 2015 |  |  | 2016 |  |  | 2017 |  |  | 2018 |  |  |
| --- | --- | --- | --- | --- | --- | --- | --- | --- | --- | --- | --- | --- | --- | --- | --- | --- | --- | --- | --- | --- | --- | --- | --- | --- |
| Prefecture | Case | Outcome |  | Case | Outcome |  | Case | Outcome |  | Case | Outcome |  | Case | Outcome |  | Case | Outcome |  | Case | Outcome |  | Case | Outcome |  |
| 01 | 1225 | 38 | 3.1% | 1299 | 37 | 2.8% | 1264 | 26 | 2.1% | 1288 | 28 | 2.2% | 1314 | 33 | 2.5% | 1209 | 39 | 3.2% | 1158 | 27 | 2.3% | 1164 | 35 | 3.0% |
| 02 | 272 | - | - | 258 | - | - | 241 | - | - | 232 | - | - | 265 | 10 | 3.8% | 226 | - | - | 209 | - | - | 171 | - | - |
| 03 | 257 | - | - | 259 | - | - | 241 | - | - | 202 | - | - | 182 | - | - | 176 | - | - | 176 | 11 | 6.3% | 163 | - | - |
| 04 | 197 | - | - | 194 | - | - | 152 | 10 | 6.6% | 194 | 11 | 5.7% | 274 | - | - | 275 | 11 | 4.0% | 264 | - | - | 250 | - | - |
| 05 | 136 | - | - | 117 | - | - | 172 | 11 | 6.4% | 202 | 10 | 5.0% | 178 | - | - | 167 | - | - | 165 | - | - | 145 | - | - |
| 06 | 175 | - | - | 182 | - | - | 156 | - | - | 179 | - | - | 187 | - | - | 157 | - | - | 162 | - | - | 135 | - | - |
| 07 | 263 | 22 | 8.4% | 230 | 17 | 7.4% | 219 | 21 | 9.6% | 207 | 12 | 5.8% | 185 | 13 | 7.0% | 194 | - | - | 204 | 17 | 8.3% | 192 | 14 | 7.3% |
| 08 | 332 | 26 | 7.8% | 433 | 24 | 5.5% | 403 | 20 | 5.0% | 433 | 26 | 6.0% | 456 | 21 | 4.6% | 419 | 15 | 3.6% | 367 | 15 | 4.1% | 405 | 22 | 5.4% |
| 09 | 244 | - | - | 281 | 10 | 3.6% | 290 | 11 | 3.8% | 265 | - | - | 267 | 16 | 6.0% | 250 | - | - | 263 | 13 | 4.9% | 283 | 12 | 4.2% |
| 10 | 252 | 20 | 7.9% | 262 | 10 | 3.8% | 218 | 13 | 6.0% | 272 | 20 | 7.4% | 260 | 19 | 7.3% | 228 | 11 | 4.8% | 210 | - | - | 218 | 14 | 6.4% |
| 11 | 634 | 36 | 5.7% | 683 | 39 | 5.7% | 660 | 35 | 5.3% | 705 | 30 | 4.3% | 645 | 37 | 5.7% | 726 | 42 | 5.8% | 666 | 43 | 6.5% | 698 | 40 | 5.7% |
| 12 | 572 | 31 | 5.4% | 585 | 36 | 6.2% | 592 | 25 | 4.2% | 590 | 18 | 3.1% | 543 | 33 | 6.1% | 540 | 24 | 4.4% | 563 | 18 | 3.2% | 564 | 29 | 5.1% |
| 13 | 1626 | 84 | 5.2% | 1646 | 74 | 4.5% | 1636 | 63 | 3.9% | 1527 | 79 | 5.2% | 1538 | 70 | 4.6% | 1406 | 66 | 4.7% | 1394 | 57 | 4.1% | 1352 | 55 | 4.1% |
| 14 | 1036 | 67 | 6.5% | 989 | 57 | 5.8% | 880 | 48 | 5.5% | 1007 | 50 | 5.0% | 962 | 51 | 5.3% | 986 | 49 | 5.0% | 999 | 42 | 4.2% | 957 | 32 | 3.3% |
| 15 | 219 | - | - | 241 | 10 | 4.1% | 224 | - | - | 220 | 12 | 5.5% | 198 | - | - | 228 | 12 | 5.3% | 229 | 10 | 4.4% | 202 | - | - |
| 16 | 168 | - | - | 131 | - | - | 155 | 13 | 8.4% | 139 | - | - | 127 | - | - | 123 | - | - | 124 | - | - | 137 | - | - |
| 17 | 130 | - | - | 114 | - | - | 112 | - | - | 104 | - | - | 105 | - | - | 104 | - | - | 106 | - | - | 77 | - | - |
| 18 | 94 | - | - | 92 | - | - | 93 | - | - | 91 | - | - | 71 | - | - | 72 | - | - | 59 | - | - | 65 | - | - |
| 19 | 75 | - | - | 76 | - | - | 96 | - | - | 80 | - | - | 83 | - | - | 82 | - | - | 68 | - | - | 74 | - | - |
| 20 | 375 | 16 | 4.3% | 445 | 25 | 5.6% | 427 | 12 | 2.8% | 435 | 18 | 4.1% | 392 | 17 | 4.3% | 362 | 11 | 3.0% | 329 | 11 | 3.3% | 342 | 15 | 4.4% |
| 21 | 212 | 13 | 6.1% | 235 | 14 | 6.0% | 210 | - | - | 179 | 16 | 8.9% | 183 | - | - | 155 | - | - | 157 | 11 | 7.0% | 196 | - | - |
| 22 | 589 | 45 | 7.6% | 548 | 40 | 7.3% | 503 | 38 | 7.6% | 494 | 30 | 6.1% | 501 | 30 | 6.0% | 471 | 20 | 4.2% | 474 | 27 | 5.7% | 437 | 18 | 4.1% |
| 23 | 889 | 43 | 4.8% | 852 | 40 | 4.7% | 854 | 31 | 3.6% | 840 | 32 | 3.8% | 891 | 25 | 2.8% | 816 | 31 | 3.8% | 847 | 25 | 3.0% | 790 | 26 | 3.3% |
| 24 | 218 | 20 | 9.2% | 241 | 13 | 5.4% | 235 | 10 | 4.3% | 230 | 10 | 4.3% | 184 | 13 | 7.1% | 208 | 10 | 4.8% | 196 | 11 | 5.6% | 199 | - | - |
| 25 | 149 | 12 | 8.1% | 127 | - | - | 150 | 10 | 6.7% | 131 | - | - | 130 | - | - | 126 | - | - | 127 | - | - | 124 | - | - |
| 26 | 258 | 16 | 6.2% | 259 | 10 | 3.9% | 252 | 11 | 4.4% | 231 | 16 | 6.9% | 237 | 13 | 5.5% | 216 | 13 | 6.0% | 206 | 11 | 5.3% | 152 | - | - |
| 27 | 1221 | 75 | 6.1% | 1249 | 69 | 5.5% | 1205 | 46 | 3.8% | 1156 | 60 | 5.2% | 1081 | 50 | 4.6% | 1057 | 50 | 4.7% | 1012 | 41 | 4.1% | 870 | 41 | 4.7% |
| 28 | 733 | 35 | 4.8% | 780 | 41 | 5.3% | 825 | 44 | 5.3% | 801 | 39 | 4.9% | 733 | 33 | 4.5% | 751 | 36 | 4.8% | 704 | 30 | 4.3% | 682 | 23 | 3.4% |
| 29 | 198 | 12 | 6.1% | 202 | 17 | 8.4% | 181 | 15 | 8.3% | 212 | 11 | 5.2% | 225 | 13 | 5.8% | 234 | 11 | 4.7% | 231 | 10 | 4.3% | 228 | 13 | 5.7% |
| 30 | 120 | - | - | 98 | - | - | 117 | - | - | 116 | - | - | 96 | - | - | 100 | 10 | 10.0% | 70 | - | - | 76 | - | - |
| 31 | 97 | - | - | 103 | - | - | 81 | - | - | 70 | - | - | 76 | - | - | 52 | - | - | 74 | - | - | 59 | - | - |
| 32 | 93 | - | - | 83 | - | - | 93 | - | - | 83 | - | - | 84 | - | - | 74 | - | - | 75 | - | - | 73 | - | - |
| 33 | 244 | 15 | 6.1% | 242 | - | - | 259 | - | - | 222 | - | - | 208 | - | - | 207 | - | - | 175 | - | - | 171 | - | - |
| 34 | 339 | 16 | 4.7% | 299 | 11 | 3.7% | 324 | 15 | 4.6% | 291 | 12 | 4.1% | 293 | 13 | 4.4% | 291 | - | - | 305 | 11 | 3.6% | 353 | 10 | 2.8% |
| 35 | 191 | 10 | 5.2% | 172 | - | - | 206 | 10 | 4.9% | 203 | 11 | 5.4% | 193 | - | - | 189 | - | - | 153 | - | - | 142 | - | - |
| 36 | 105 | - | - | 93 | - | - | 103 | - | - | 93 | - | - | 70 | - | - | 60 | - | - | 61 | - | - | 73 | - | - |
| 37 | 84 | - | - | 83 | - | - | 84 | - | - | 84 | - | - | 108 | - | - | 74 | - | - | 66 | - | - | 68 | - | - |
| 38 | 135 | - | - | 135 | - | - | 110 | - | - | 177 | - | - | 127 | - | - | 155 | - | - | 165 | 13 | 7.9% | 125 | - | - |
| 39 | 120 | - | - | 96 | - | - | 120 | - | - | 82 | - | - | 83 | - | - | 96 | - | - | 104 | - | - | 94 | - | - |
| 40 | 803 | 34 | 4.2% | 803 | 36 | 4.5% | 711 | 28 | 3.9% | 768 | 22 | 2.9% | 723 | 18 | 2.5% | 621 | 16 | 2.6% | 624 | 26 | 4.2% | 616 | 14 | 2.3% |
| 41 | 102 | - | - | 99 | - | - | 104 | - | - | 136 | - | - | 118 | - | - | 118 | - | - | 115 | - | - | 102 | - | - |
| 42 | 229 | - | - | 207 | - | - | 192 | - | - | 178 | - | - | 185 | - | - | 198 | - | - | 195 | - | - | 202 | - | - |
| 43 | 343 | - | - | 316 | 11 | 3.5% | 306 | - | - | 330 | - | - | 287 | 13 | 4.5% | 265 | - | - | 293 | - | - | 242 | - | - |
| 44 | 106 | - | - | 115 | - | - | 89 | - | - | 73 | - | - | 101 | - | - | 118 | - | - | 90 | - | - | 85 | - | - |
| 45 | 146 | 11 | 7.5% | 168 | - | - | 145 | - | - | 108 | - | - | 151 | - | - | 170 | - | - | 151 | - | - | 143 | - | - |
| 46 | 301 | - | - | 290 | 10 | 3.4% | 296 | 16 | 5.4% | 292 | 14 | 4.8% | 290 | - | - | 248 | 17 | 6.9% | 242 | - | - | 242 | 10 | 4.1% |
| 47 | 144 | - | - | 182 | - | - | 124 | - | - | 121 | - | - | 127 | - | - | 130 | - | - | 151 | - | - | 134 | - | - |

- indicates <10.

**Fig. 2.**
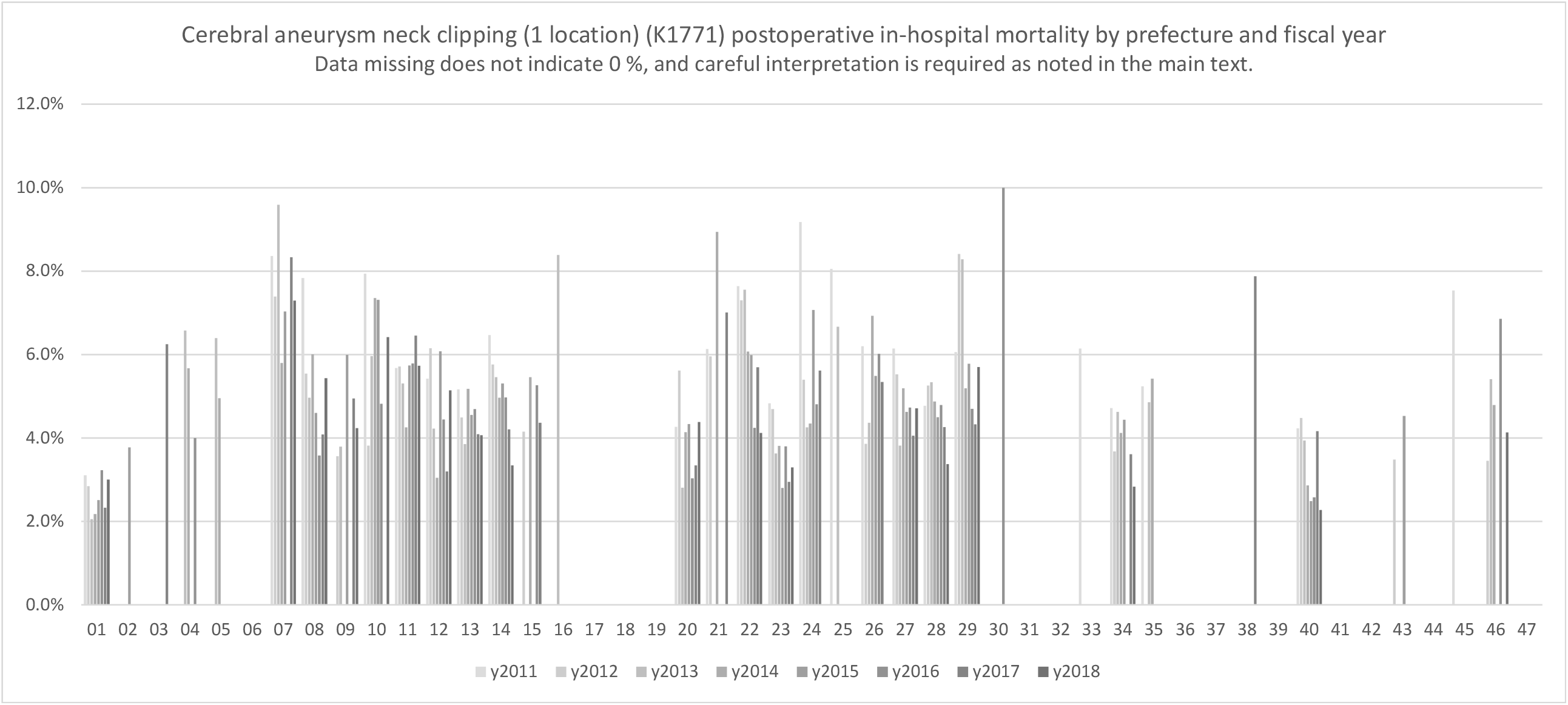
Cerebral aneurysm neck clipping (one location) (K1771)

### Coronary artery and aortic bypass grafting (K5521, K5522, K552-21, and K552-22)

Tables 3-1, 3-2, 3-3, and 3-4 show the results for coronary artery and aortic bypass grafting with one anastomosis with the heart–lung machine (K5521), more than two anastomoses with the heart–lung machine(K5522), with one anastomosis without the heart–lung machine(K552-21), and more than two anastomoses without the heart–lung machine (K552-22). Of the four, Table 3-2 provides relatively more numeric results, but in the others, numeric results were available for the denominator, and the number of numerators in many areas was too small to be tabulated. Figure 3 graphically depicts the postoperative in-hospital mortality in coronary artery and aortic bypass grafting of more than two anastomoses (K5522, Table 3-2). Although only a few areas can be depicted as a result of the tally, regional differences were noted in this tally. On the contrary, coronary artery bypass surgery is classified into four reimbursement categories, as shown in four tables. These can certainly be considered different surgeries; however, the numerical value of outcomes for other coronary artery bypass grafting could not be depicted since the number of each outcome was small.

**Table 3-1.**
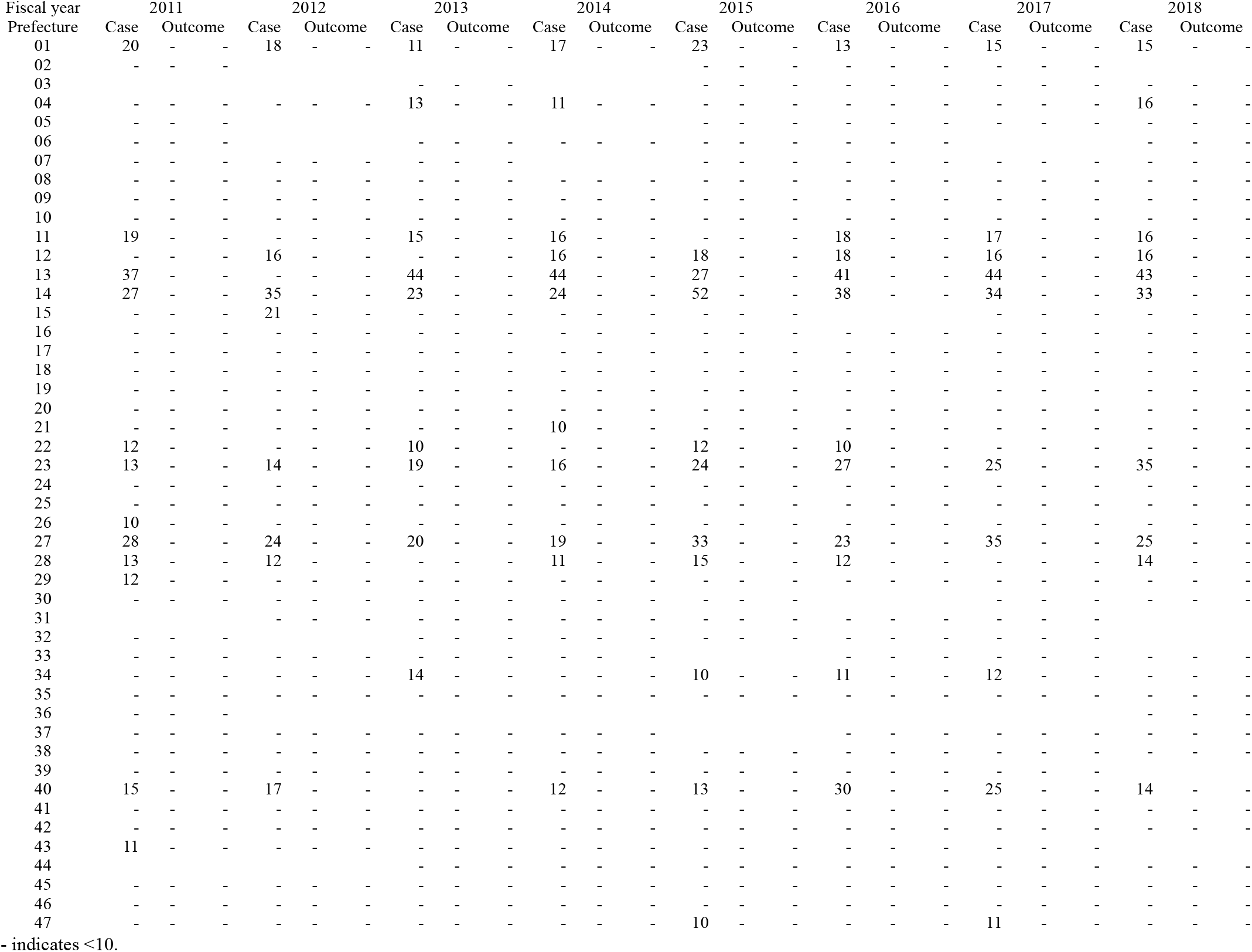
Coronary artery and aortic bypass grafting with one anastomosis using a heart–lung machine (K5521), case number, and outcomes of postoperative in-hospital mortality

**Table 3-2.**
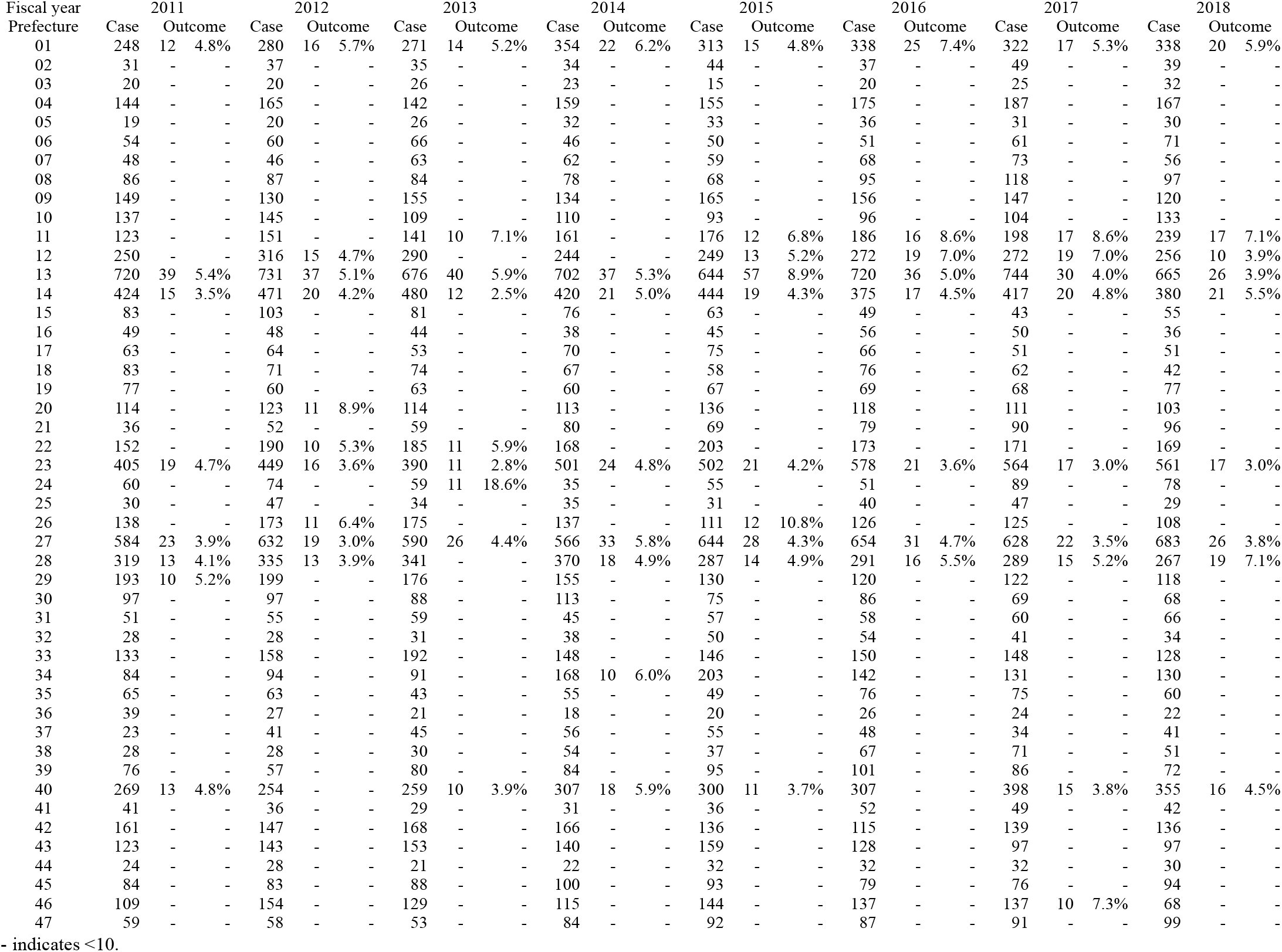
Coronary artery and aortic bypass grafting with more than two anastomoses using a heart–lung machine (K5522), case number, and outcomes of postoperative in-hospital mortality

| Fiscal year | 2011 |  |  | 2012 |  |  | 2013 |  |  | 2014 |  |  | 2015 |  |  | 2016 |  |  | 2017 |  |  | 2018 |  |  |
| --- | --- | --- | --- | --- | --- | --- | --- | --- | --- | --- | --- | --- | --- | --- | --- | --- | --- | --- | --- | --- | --- | --- | --- | --- |
| Prefecture | Case | Outcome |  | Case | Outcome |  | Case | Outcome |  | Case | Outcome |  | Case | Outcome |  | Case | Outcome |  | Case | Outcome |  | Case | Outcome |  |
| 01 | 248 | 12 | 4.8% | 280 | 16 | 5.7% | 271 | 14 | 5.2% | 354 | 22 | 6.2% | 313 | 15 | 4.8% | 338 | 25 | 7.4% | 322 | 17 | 5.3% | 338 | 20 | 5.9% |
| 02 | 31 | - | - | 37 | - | - | 35 | - | - | 34 | - | - | 44 | - | - | 37 | - | - | 49 | - | - | 39 | - | - |
| 03 | 20 | - | - | 20 | - | - | 26 | - | - | 23 | - | - | 15 | - | - | 20 | - | - | 25 | - | - | 32 | - | - |
| 04 | 144 | - | - | 165 | - | - | 142 | - | - | 159 | - | - | 155 | - | - | 175 | - | - | 187 | - | - | 167 | - | - |
| 05 | 19 | - | - | 20 | - | - | 26 | - | - | 32 | - | - | 33 | - | - | 36 | - | - | 31 | - | - | 30 | - | - |
| 06 | 54 | - | - | 60 | - | - | 66 | - | - | 46 | - | - | 50 | - | - | 51 | - | - | 61 | - | - | 71 | - | - |
| 07 | 48 | - | - | 46 | - | - | 63 | - | - | 62 | - | - | 59 | - | - | 68 | - | - | 73 | - | - | 56 | - | - |
| 08 | 86 | - | - | 87 | - | - | 84 | - | - | 78 | - | - | 68 | - | - | 95 | - | - | 118 | - | - | 97 | - | - |
| 09 | 149 | - | - | 130 | - | - | 155 | - | - | 134 | - | - | 165 | - | - | 156 | - | - | 147 | - | - | 120 | - | - |
| 10 | 137 | - | - | 145 | - | - | 109 | - | - | 110 | - | - | 93 | - | - | 96 | - | - | 104 | - | - | 133 | - | - |
| 11 | 123 | - | - | 151 | - | - | 141 | 10 | 7.1% | 161 | - | - | 176 | 12 | 6.8% | 186 | 16 | 8.6% | 198 | 17 | 8.6% | 239 | 17 | 7.1% |
| 12 | 250 | - | - | 316 | 15 | 4.7% | 290 | - | - | 244 | - | - | 249 | 13 | 5.2% | 272 | 19 | 7.0% | 272 | 19 | 7.0% | 256 | 10 | 3.9% |
| 13 | 720 | 39 | 5.4% | 731 | 37 | 5.1% | 676 | 40 | 5.9% | 702 | 37 | 5.3% | 644 | 57 | 8.9% | 720 | 36 | 5.0% | 744 | 30 | 4.0% | 665 | 26 | 3.9% |
| 14 | 424 | 15 | 3.5% | 471 | 20 | 4.2% | 480 | 12 | 2.5% | 420 | 21 | 5.0% | 444 | 19 | 4.3% | 375 | 17 | 4.5% | 417 | 20 | 4.8% | 380 | 21 | 5.5% |
| 15 | 83 | - | - | 103 | - | - | 81 | - | - | 76 | - | - | 63 | - | - | 49 | - | - | 43 | - | - | 55 | - | - |
| 16 | 49 | - | - | 48 | - | - | 44 | - | - | 38 | - | - | 45 | - | - | 56 | - | - | 50 | - | - | 36 | - | - |
| 17 | 63 | - | - | 64 | - | - | 53 | - | - | 70 | - | - | 75 | - | - | 66 | - | - | 51 | - | - | 51 | - | - |
| 18 | 83 | - | - | 71 | - | - | 74 | - | - | 67 | - | - | 58 | - | - | 76 | - | - | 62 | - | - | 42 | - | - |
| 19 | 77 | - | - | 60 | - | - | 63 | - | - | 60 | - | - | 67 | - | - | 69 | - | - | 68 | - | - | 77 | - | - |
| 20 | 114 | - | - | 123 | 11 | 8.9% | 114 | - | - | 113 | - | - | 136 | - | - | 118 | - | - | 111 | - | - | 103 | - | - |
| 21 | 36 | - | - | 52 | - | - | 59 | - | - | 80 | - | - | 69 | - | - | 79 | - | - | 90 | - | - | 96 | - | - |
| 22 | 152 | - | - | 190 | 10 | 5.3% | 185 | 11 | 5.9% | 168 | - | - | 203 | - | - | 173 | - | - | 171 | - | - | 169 | - | - |
| 23 | 405 | 19 | 4.7% | 449 | 16 | 3.6% | 390 | 11 | 2.8% | 501 | 24 | 4.8% | 502 | 21 | 4.2% | 578 | 21 | 3.6% | 564 | 17 | 3.0% | 561 | 17 | 3.0% |
| 24 | 60 | - | - | 74 | - | - | 59 | 11 | 18.6% | 35 | - | - | 55 | - | - | 51 | - | - | 89 | - | - | 78 | - | - |
| 25 | 30 | - | - | 47 | - | - | 34 | - | - | 35 | - | - | 31 | - | - | 40 | - | - | 47 | - | - | 29 | - | - |
| 26 | 138 | - | - | 173 | 11 | 6.4% | 175 | - | - | 137 | - | - | 111 | 12 | 10.8% | 126 | - | - | 125 | - | - | 108 | - | - |
| 27 | 584 | 23 | 3.9% | 632 | 19 | 3.0% | 590 | 26 | 4.4% | 566 | 33 | 5.8% | 644 | 28 | 4.3% | 654 | 31 | 4.7% | 628 | 22 | 3.5% | 683 | 26 | 3.8% |
| 28 | 319 | 13 | 4.1% | 335 | 13 | 3.9% | 341 | - | - | 370 | 18 | 4.9% | 287 | 14 | 4.9% | 291 | 16 | 5.5% | 289 | 15 | 5.2% | 267 | 19 | 7.1% |
| 29 | 193 | 10 | 5.2% | 199 | - | - | 176 | - | - | 155 | - | - | 130 | - | - | 120 | - | - | 122 | - | - | 118 | - | - |
| 30 | 97 | - | - | 97 | - | - | 88 | - | - | 113 | - | - | 75 | - | - | 86 | - | - | 69 | - | - | 68 | - | - |
| 31 | 51 | - | - | 55 | - | - | 59 | - | - | 45 | - | - | 57 | - | - | 58 | - | - | 60 | - | - | 66 | - | - |
| 32 | 28 | - | - | 28 | - | - | 31 | - | - | 38 | - | - | 50 | - | - | 54 | - | - | 41 | - | - | 34 | - | - |
| 33 | 133 | - | - | 158 | - | - | 192 | - | - | 148 | - | - | 146 | - | - | 150 | - | - | 148 | - | - | 128 | - | - |
| 34 | 84 | - | - | 94 | - | - | 91 | - | - | 168 | 10 | 6.0% | 203 | - | - | 142 | - | - | 131 | - | - | 130 | - | - |
| 35 | 65 | - | - | 63 | - | - | 43 | - | - | 55 | - | - | 49 | - | - | 76 | - | - | 75 | - | - | 60 | - | - |
| 36 | 39 | - | - | 27 | - | - | 21 | - | - | 18 | - | - | 20 | - | - | 26 | - | - | 24 | - | - | 22 | - | - |
| 37 | 23 | - | - | 41 | - | - | 45 | - | - | 56 | - | - | 55 | - | - | 48 | - | - | 34 | - | - | 41 | - | - |
| 38 | 28 | - | - | 28 | - | - | 30 | - | - | 54 | - | - | 37 | - | - | 67 | - | - | 71 | - | - | 51 | - | - |
| 39 | 76 | - | - | 57 | - | - | 80 | - | - | 84 | - | - | 95 | - | - | 101 | - | - | 86 | - | - | 72 | - | - |
| 40 | 269 | 13 | 4.8% | 254 | - | - | 259 | 10 | 3.9% | 307 | 18 | 5.9% | 300 | 11 | 3.7% | 307 | - | - | 398 | 15 | 3.8% | 355 | 16 | 4.5% |
| 41 | 41 | - | - | 36 | - | - | 29 | - | - | 31 | - | - | 36 | - | - | 52 | - | - | 49 | - | - | 42 | - | - |
| 42 | 161 | - | - | 147 | - | - | 168 | - | - | 166 | - | - | 136 | - | - | 115 | - | - | 139 | - | - | 136 | - | - |
| 43 | 123 | - | - | 143 | - | - | 153 | - | - | 140 | - | - | 159 | - | - | 128 | - | - | 97 | - | - | 97 | - | - |
| 44 | 24 | - | - | 28 | - | - | 21 | - | - | 22 | - | - | 32 | - | - | 32 | - | - | 32 | - | - | 30 | - | - |
| 45 | 84 | - | - | 83 | - | - | 88 | - | - | 100 | - | - | 93 | - | - | 79 | - | - | 76 | - | - | 94 | - | - |
| 46 | 109 | - | - | 154 | - | - | 129 | - | - | 115 | - | - | 144 | - | - | 137 | - | - | 137 | 10 | 7.3% | 68 | - | - |
| 47 | 59 | - | - | 58 | - | - | 53 | - | - | 84 | - | - | 92 | - | - | 87 | - | - | 91 | - | - | 99 | - | - |

- indicates <10.

**Table 3-3.** Coronary artery and aortic bypass grafting with one anastomosis without a heart–lung machine (K552-21), case number, and outcomes of postoperative in-hospital mortality

| Fiscal year | 2011 |  | 2012 |  | 2013 |  | 2014 |  | 2015 |  | 2016 |  | 2017 |  | 2018 |  |
| --- | --- | --- | --- | --- | --- | --- | --- | --- | --- | --- | --- | --- | --- | --- | --- | --- |
| Prefecture | Case | Outcome | Case | Outcome | Case | Outcome | Case | Outcome | Case | Outcome | Case | Outcome | Case | Outcome | Case | Outcome |
| 01 | 46 | - | 60 | - | 60 | - | 51 | - | 42 | - | 84 | - | 83 | - | 52 | - |
| 02 | 15 | - | 13 | - | 21 | - | 25 | - | 11 | - | 15 | - | 13 | - | - | - |
| 03 | 10 | - | 15 | - | - | - | - | - | 13 | - | - | - | 16 | - | 14 | - |
| 04 | 13 | - | 11 | - | 13 | - | 11 | - | 11 | - | 14 | - | 11 | - | 18 | - |
| 05 | 14 | - | - | - | - | - | - | - | - | - | - | - | - | - | - | - |
| 06 | 20 | - | 14 | - | - | - | 12 | - | 12 | - | - | - | - | - | 12 | - |
| 07 | - | - | 13 | - | - | - | - | - | 10 | - | - | - | - | - | - | - |
| 08 | - | - | - | - | 15 | - | 13 | - | - | - | 15 | - | 12 | - | - | - |
| 09 | - | - | - | - | - | - | - | - | - | - | - | - | - | - | - | - |
| 10 | 19 | - | 18 | - | 14 | - | 13 | - | 10 | - | 15 | - | - | - | - | - |
| 11 | 20 | - | 19 | - | 19 | - | 26 | - | 31 | - | 22 | - | 23 | - | 29 | - |
| 12 | 33 | - | 42 | - | 24 | - | 25 | - | 34 | - | 36 | - | 60 | - | 79 | - |
| 13 | 68 | - | 100 | - | 90 | - | 104 | - | 102 | - | 73 | - | 95 | - | 107 | - |
| 14 | 59 | - | 52 | - | 51 | - | 59 | - | 52 | - | 66 | - | 73 | - | 78 | - |
| 15 | - | - | - | - | - | - | - | - | - | - | - | - | - | - | - | - |
| 16 | - | - | - | - | - | - | - | - | - | - | 10 | - | - | - | - | - |
| 17 | - | - | 10 | - | - | - | - | - | - | - | - | - | - | - | - | - |
| 18 | - | - | - | - | - | - | - | - | - | - | - | - | - | - | - | - |
| 19 | - | - | - | - | - | - | - | - | - | - | - | - | - | - | - | - |
| 20 | 20 | - | 29 | - | 25 | - | 24 | - | 19 | - | 17 | - | 25 | - | 24 | - |
| 21 | 17 | - | 10 | - | 14 | - | - | - | 14 | - | 16 | - | - | - | 20 | - |
| 22 | 11 | - | 13 | - | 30 | - | 29 | - | 32 | - | 30 | - | 26 | - | 22 | - |
| 23 | 66 | - | 60 | - | 76 | - | 54 | - | 72 | - | 70 | - | 59 | - | 46 | - |
| 24 | - | - | - | - | - | - | - | - | 14 | - | - | - | 11 | - | 15 | - |
| 25 | - | - | - | - | - | - | - | - | 12 | - | - | - | - | - | - | - |
| 26 | 12 | - | 18 | - | 13 | - | 10 | - | - | - | 13 | - | 14 | - | - | - |
| 27 | 38 | - | 66 | - | 42 | - | 39 | - | 46 | - | 36 | - | 42 | - | 40 | - |
| 28 | 15 | - | 21 | - | 20 | - | 34 | - | 19 | - | 23 | - | 18 | - | 31 | - |
| 29 | - | - | - | - | - | - | - | - | - | - | - | - | - | - | - | - |
| 30 | - | - | - | - | - | - | - | - | - | - | 12 | - | - | - | - | - |
| 31 | - | - | - | - | - | - | - | - | - | - | - | - | - | - | - | - |
| 32 | - | - | - | - | - | - | - | - | - | - | - | - | - | - | 10 | - |
| 33 | 13 | - | 18 | - | 13 | - | 19 | - | 21 | - | 22 | - | 11 | - | 14 | - |
| 34 | 18 | - | 20 | - | 18 | - | 14 | - | 15 | - | 18 | - | 16 | - | 16 | - |
| 35 | - | - | 18 | - | 14 | - | 20 | - | 10 | - | 18 | - | - | - | - | - |
| 36 | - | - | - | - | - | - | - | - | - | - | - | - | - | - | - | - |
| 37 | - | - | 13 | - | - | - | 11 | - | - | - | - | - | - | - | - | - |
| 38 | - | - | 10 | - | - | - | 11 | - | 14 | - | - | - | - | - | 11 | - |
| 39 | - | - | - | - | - | - | - | - | - | - | - | - | - | - | - | - |
| 40 | 37 | - | 21 | - | 20 | - | 19 | - | 30 | - | 18 | - | 18 | - | 14 | - |
| 41 | - | - | - | - | - | - | - | - | - | - | - | - | - | - | - | - |
| 42 | 12 | - | 11 | - | - | - | - | - | - | - | - | - | - | - | - | - |
| 43 | - | - | 13 | - | - | - | 11 | - | - | - | - | - | - | - | - | - |
| 44 | - | - | - | - | - | - | - | - | - | - | - | - | 14 | - | 19 | - |
| 45 | - | - | - | - | - | - | - | - | - | - | - | - | - | - | - | - |
| 46 | - | - | - | - | - | - | - | - | 15 | - | - | - | - | - | - | - |
| 47 | - | - | 12 | - | 16 | - | 19 | - | 13 | - | 11 | - | 12 | - | - | - |

- indicates less than 10.

**Table 3-4.** Coronary artery and aortic bypass grafting with more than two anastomoses without a heart–lung machine (K552-22), case number, and outcomes of postoperative in-hospital mortality

| Fiscal year | 2011 |  |  | 2012 |  |  | 2013 |  |  | 2014 |  |  | 2015 |  |  | 2016 |  |  | 2017 |  |  | 2018 |  |  |
| --- | --- | --- | --- | --- | --- | --- | --- | --- | --- | --- | --- | --- | --- | --- | --- | --- | --- | --- | --- | --- | --- | --- | --- | --- |
| Prefecture | Case | Outcome |  | Case | Outcome |  | Case | Outcome |  | Case | Outcome |  | Case | Outcome |  | Case | Outcome |  | Case | Outcome |  | Case | Outcome |  |
| 01 | 414 | 10 | 2.4% | 477 | - | - | 451 | 13 | 2.9% | 469 | - | - | 562 | - | - | 528 | - | - | 545 | - | - | 471 | 13 | 2.8% |
| 02 | 135 | - | - | 109 | - | - | 115 | - | - | 82 | - | - | 121 | - | - | 89 | - | - | 94 | - | - | 107 | - | - |
| 03 | 95 | - | - | 80 | - | - | 68 | - | - | 80 | - | - | 54 | - | - | 73 | - | - | 85 | - | - | 99 | - | - |
| 04 | 49 | - | - | 62 | - | - | 38 | - | - | 31 | - | - | 57 | - | - | 57 | - | - | 45 | - | - | 42 | - | - |
| 05 | 29 | - | - | 29 | - | - | 19 | - | - | 15 | - | - | 16 | - | - | - | - | - | 10 | - | - | 11 | - | - |
| 06 | 28 | - | - | 38 | - | - | 45 | - | - | 55 | - | - | 68 | - | - | 37 | - | - | 42 | - | - | 58 | - | - |
| 07 | 71 | - | - | 76 | - | - | 68 | - | - | 75 | - | - | 80 | - | - | 88 | - | - | 53 | - | - | 67 | - | - |
| 08 | 103 | - | - | 94 | - | - | 103 | - | - | 94 | - | - | 105 | - | - | 115 | - | - | 96 | - | - | 121 | - | - |
| 09 | 20 | - | - | 18 | - | - | 34 | - | - | 26 | - | - | 12 | - | - | 20 | - | - | 24 | - | - | 19 | - | - |
| 10 | 52 | - | - | 78 | - | - | 97 | - | - | 66 | - | - | 72 | - | - | 43 | - | - | 34 | - | - | 33 | - | - |
| 11 | 361 | - | - | 354 | - | - | 351 | - | - | 312 | - | - | 343 | - | - | 364 | - | - | 350 | - | - | 361 | - | - |
| 12 | 183 | - | - | 215 | - | - | 272 | - | - | 359 | - | - | 363 | - | - | 379 | - | - | 345 | - | - | 366 | - | - |
| 13 | 1024 | 14 | 1.4% | 1178 | 10 | 0.8% | 1148 | 21 | 1.8% | 1168 | 13 | 1.1% | 1144 | 15 | 1.3% | 1048 | 14 | 1.3% | 1083 | 15 | 1.4% | 1083 | - | - |
| 14 | 339 | - | - | 331 | - | - | 318 | - | - | 391 | - | - | 475 | - | - | 503 | 10 | 2.0% | 425 | - | - | 424 | - | - |
| 15 | 84 | - | - | 99 | - | - | 82 | - | - | 115 | - | - | 90 | - | - | 67 | - | - | 70 | - | - | 49 | - | - |
| 16 | 88 | - | - | 79 | - | - | 94 | - | - | 102 | - | - | 110 | - | - | 83 | - | - | 63 | - | - | 92 | - | - |
| 17 | 84 | - | - | 85 | - | - | 75 | - | - | 44 | - | - | 70 | - | - | 89 | - | - | 100 | - | - | 98 | - | - |
| 18 | 29 | - | - | 38 | - | - | 46 | - | - | 28 | - | - | 48 | - | - | 30 | - | - | - | - | - | 38 | - | - |
| 19 | - | - | - | - | - | - | - | - | - | - | - | - | - | - | - | - | - | - | 37 | - | - | - | - | - |
| 20 | 122 | - | - | 129 | - | - | 137 | - | - | 132 | - | - | 134 | - | - | 123 | - | - | 99 | - | - | 113 | - | - |
| 21 | 40 | - | - | 70 | - | - | 42 | - | - | 43 | - | - | 86 | - | - | 117 | - | - | 119 | - | - | 137 | - | - |
| 22 | 218 | - | - | 256 | - | - | 236 | - | - | 235 | - | - | 219 | 10 | 4.6% | 223 | - | - | 218 | - | - | 226 | - | - |
| 23 | 466 | - | - | 512 | - | - | 547 | - | - | 496 | - | - | 490 | - | - | 495 | - | - | 502 | - | - | 518 | 11 | 2.1% |
| 24 | 151 | - | - | 166 | - | - | 154 | - | - | 166 | - | - | 143 | - | - | 127 | - | - | 116 | - | - | 126 | - | - |
| 25 | 139 | - | - | 136 | - | - | 130 | - | - | 159 | - | - | 131 | - | - | 125 | - | - | 129 | - | - | 116 | - | - |
| 26 | 154 | - | - | 156 | - | - | 129 | - | - | 120 | - | - | 144 | - | - | 127 | - | - | 133 | - | - | 140 | - | - |
| 27 | 443 | - | - | 492 | - | - | 507 | 10 | 2.0% | 453 | 10 | 2.2% | 478 | - | - | 425 | - | - | 421 | - | - | 381 | - | - |
| 28 | 179 | - | - | 233 | - | - | 255 | - | - | 250 | - | - | 273 | - | - | 202 | - | - | 187 | - | - | 236 | - | - |
| 29 | 34 | - | - | 17 | - | - | 40 | - | - | 48 | - | - | 50 | - | - | 61 | - | - | 57 | - | - | 60 | - | - |
| 30 | 74 | - | - | 98 | - | - | 134 | - | - | 110 | - | - | 97 | - | - | 95 | - | - | 102 | - | - | 103 | - | - |
| 31 | 21 | - | - | 39 | - | - | 17 | - | - | 24 | - | - | 21 | - | - | 16 | - | - | 16 | - | - | 12 | - | - |
| 32 | 23 | - | - | 28 | - | - | 31 | - | - | 34 | - | - | 32 | - | - | 22 | - | - | 19 | - | - | 28 | - | - |
| 33 | 145 | - | - | 170 | - | - | 182 | - | - | 184 | - | - | 128 | - | - | 122 | - | - | 132 | - | - | 124 | - | - |
| 34 | 168 | - | - | 148 | - | - | 135 | - | - | 103 | - | - | 63 | - | - | 111 | - | - | 83 | - | - | 76 | - | - |
| 35 | 102 | - | - | 107 | - | - | 120 | - | - | 148 | - | - | 98 | - | - | 110 | - | - | 124 | - | - | 84 | - | - |
| 36 | 88 | - | - | 80 | - | - | 64 | - | - | 64 | - | - | 62 | - | - | 58 | - | - | 54 | - | - | 50 | - | - |
| 37 | 79 | - | - | 72 | - | - | 69 | - | - | 73 | - | - | 46 | - | - | 43 | - | - | 65 | - | - | 45 | - | - |
| 38 | 110 | - | - | 112 | - | - | 100 | - | - | 87 | - | - | 79 | - | - | 84 | - | - | 93 | - | - | 91 | - | - |
| 39 | 79 | - | - | 64 | - | - | 43 | - | - | 57 | - | - | 49 | - | - | 32 | - | - | 31 | - | - | 35 | - | - |
| 40 | 392 | - | - | 432 | - | - | 405 | - | - | 345 | - | - | 316 | - | - | 230 | - | - | 253 | - | - | 226 | - | - |
| 41 | 54 | - | - | 66 | - | - | 78 | - | - | 51 | - | - | 59 | - | - | 55 | - | - | 38 | - | - | 23 | - | - |
| 42 | 18 | - | - | 57 | - | - | 25 | - | - | 40 | - | - | 63 | - | - | 58 | - | - | 50 | - | - | 53 | - | - |
| 43 | 86 | - | - | 53 | - | - | 72 | - | - | 71 | - | - | 87 | - | - | 86 | - | - | 80 | - | - | 78 | - | - |
| 44 | 72 | - | - | 83 | - | - | 111 | - | - | 126 | - | - | 118 | - | - | 104 | - | - | 122 | - | - | 109 | - | - |
| 45 | 54 | - | - | 94 | - | - | 80 | - | - | 73 | - | - | 75 | - | - | 48 | - | - | 45 | - | - | 35 | - | - |
| 46 | 68 | - | - | 86 | - | - | 97 | - | - | 149 | - | - | 104 | - | - | 108 | - | - | 123 | - | - | 165 | - | - |
| 47 | 170 | - | - | 170 | - | - | 123 | - | - | 112 | - | - | 118 | - | - | 110 | - | - | 137 | - | - | 119 | - | - |

- indicates <10.

**Fig. 3.**
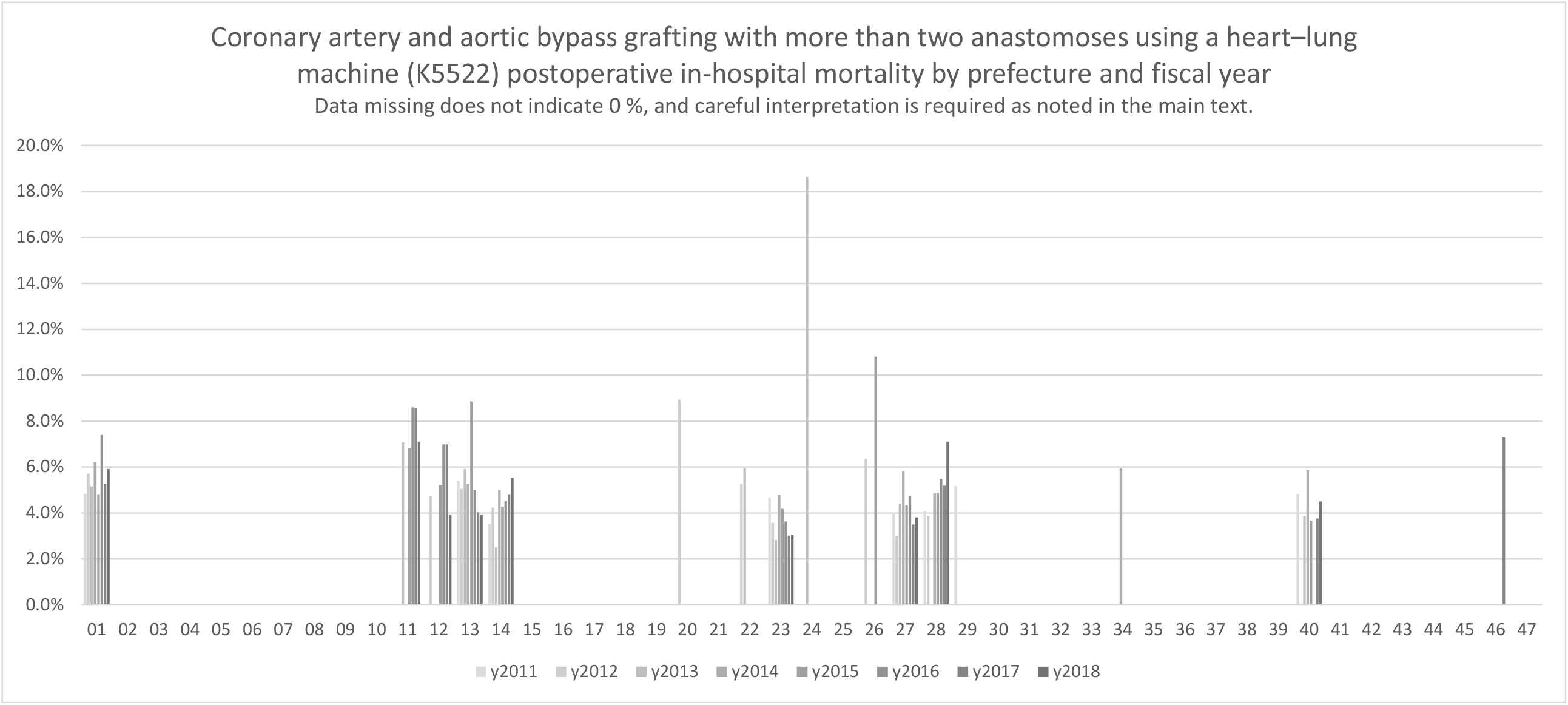
Coronary artery and aortic bypass grafting with more than two anastomoses using a heart–lung machine (K5522)

### Tracheotomy (K386)

Table 4 shows the results of tracheotomy cases. Figure 4 graphically depicts the postoperative in-hospital mortality. This tabulation provides a relatively large number of figures for both the denominator and numerator, and in many areas, comparisons can be made both across regions and over time. Although the tabulation is an analysis at the prefectural level, which means the collections widely in each region, clear regional differences are depicted in the index values; thus, the implications should be carefully interpreted as in the Discussion. The changes over time do not appear to be uniform in direction among regions, although the results for all regions combined show a decreasing trend over time.

**Table 4.**
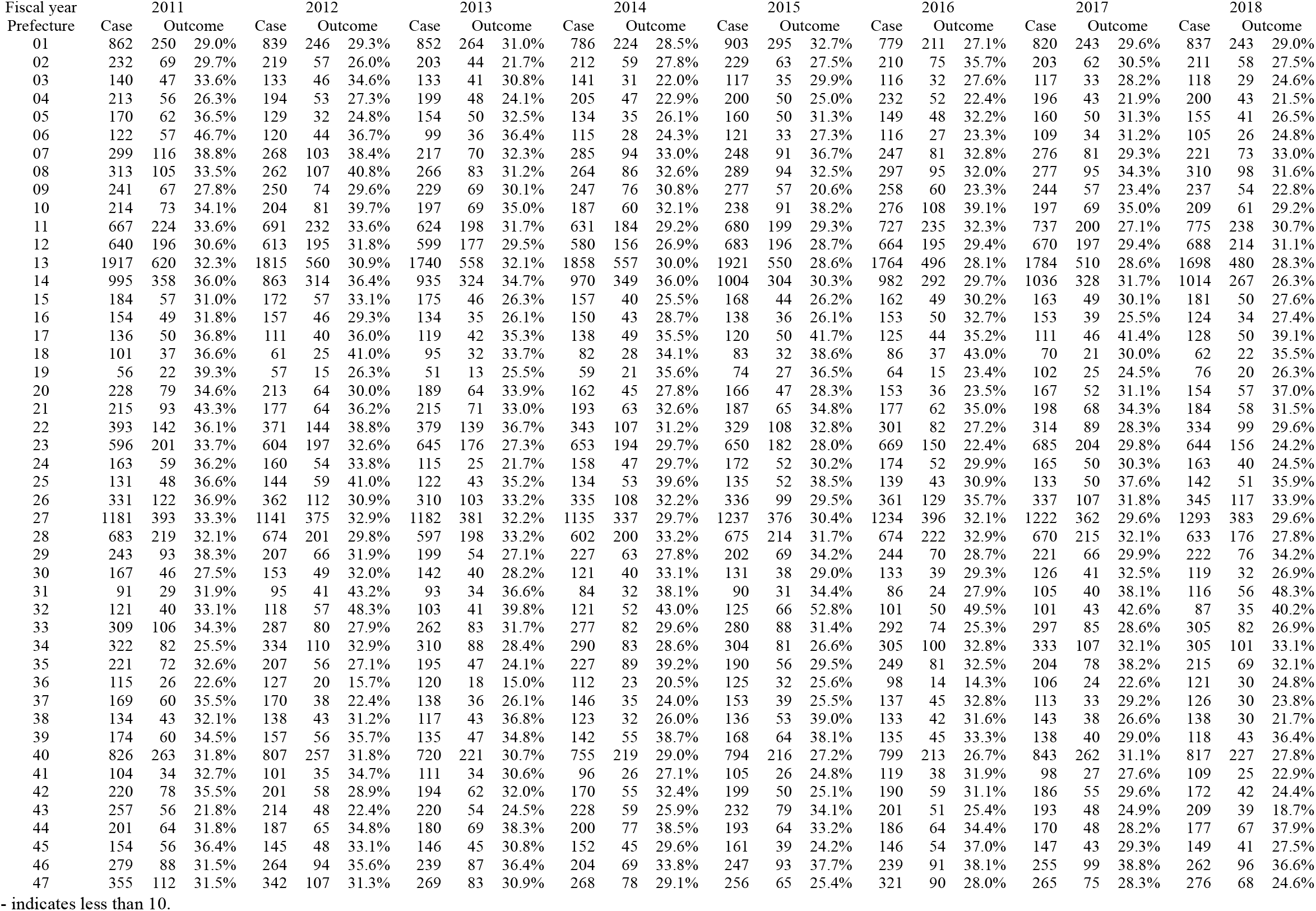
Tracheotomy (K386), case number, and outcomes of postoperative in-hospital mortality

**Fig. 4.**
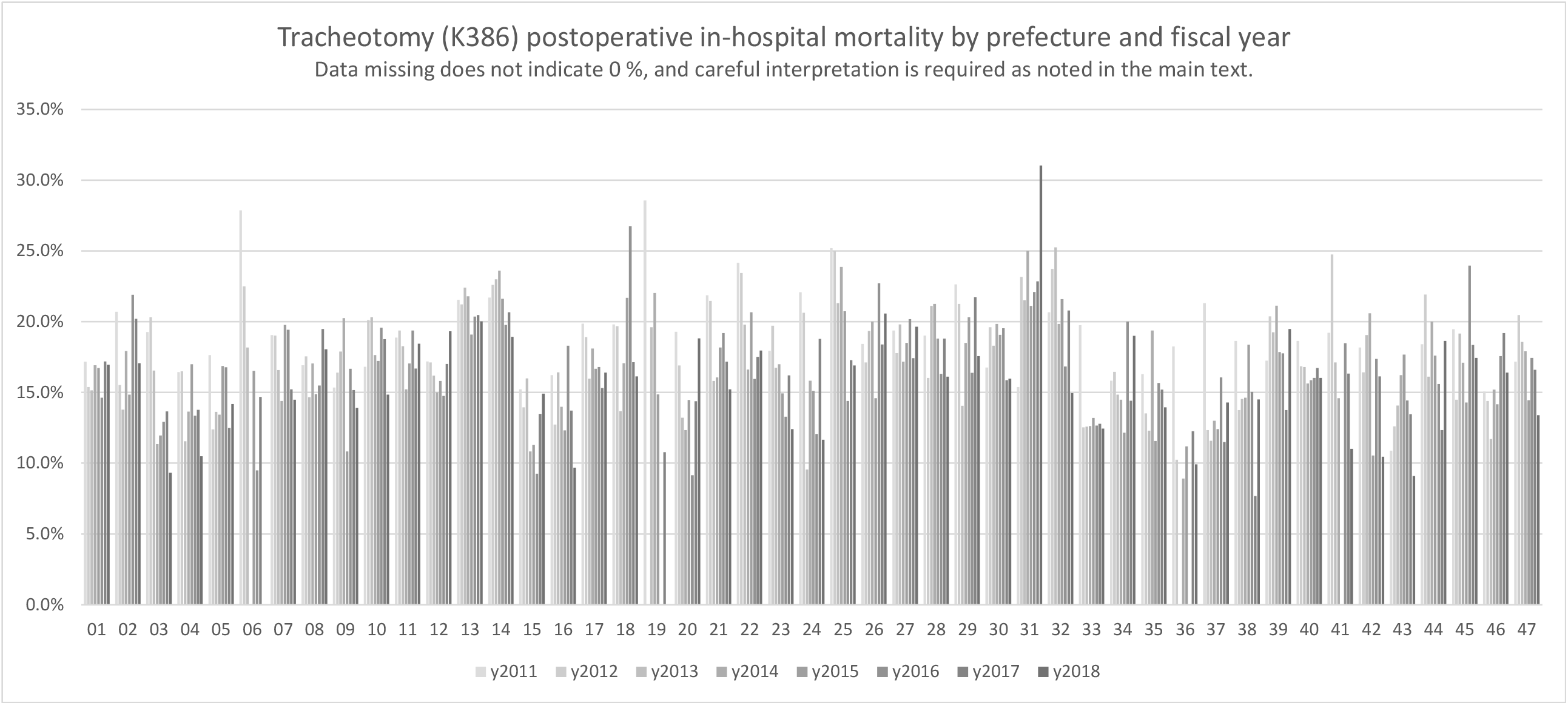
Tracheotomy (K386)

## Discussion

This study analyzed surgical outcomes at hospitals throughout Japan by region and over time using the highly comprehensive DPC database and provided comprehensive result data. Despite the simplicity of the method and broad analysis at the prefecture level, differences by region were depicted as a result of the data analysis in all surgical fields. The most important aspect in the results of this study is its comprehensiveness of dealing data from numerous hospitals. As mentioned in the Introduction, the use of the DPC database, which is now generated in large part of Japanese hospitals, makes it possible to analyze actual conditions widely that may not always be collectible by registry or other sources. This makes it possible to investigate the more actual implementation of procedures as never before. In addition, all surgeries were included in the analysis of this study, and it was possible to examine all surgeries, although the interpretation of the results varied and requires caution and all results are presented only in the supplement.

In this paper, the results presented are only some selections from all fields, but in the actual results, regional differences and changes over time are easily seen to some extent. On the contrary, negative results without noticeable differences or were unchanged can be also important for review. Although it depends on the purpose of the future research or project, the availability of data in all fields, such as in this study, is considered very valuable.

The study has several limitations and important caveats. There are many missing values. For example, if the number of cases is 300, 3% of the outcomes are only nine cases, which results in not being obtainable as an aggregate value. In addition, this study made the comparison at a rough level: the prefectural level by year, and it is essentially possible to analyze at a more detailed regional level or time span. For example, artificial head insertion (K0811) includes that for both the shoulder and hip. A separate analysis could be possible by combining disease names or using different master codes from different DPC datasets. However, a simple detailed analysis will yield fewer figures that can be obtained or presented. Even though this study made a relatively rough unit-level analysis, this can show some trends and differences, and results can be used as useful information for future studies, such as knowing fields that need to be looked at in more detail or areas that need to be collected and analyzed more thoroughly.

In this study, in-hospital mortality was selected as the outcome, but postoperative 30-day in-hospital mortality was also considered. However, the number of outcomes within 30 days is naturally smaller and could increase missing data. Furthermore, as a rule of data publication, it was necessary to avoid inference from other data with cells having <10. For example, if presenting both in-hospital mortality and in-hospital mortality within 30 days, it could be partly possible to calculate a small number of in-hospital mortality after 31 days. Therefore, only one type of outcome was used in this study, but there is the difficulty of balancing the presentation of useful data with privacy considerations.

The resulting unobtained numerical frames were somewhat expected, but somewhat more missing data were generated. For example, in coronary artery bypass surgery, the author had assumed that not a small number of outcomes could have appeared and that those actual comparisons would be possible in more areas. However, not only that the number of cases was small in some areas but also the numbers of outcomes; thus, many numerical results were unobtainable. To get around this, for example, it was possible to combine four separate coronary artery bypass procedures into one analysis or to integrate several similar procedures. It would be more useful to present both the combined aggregate and the fine-grained aggregate values; however, in principle, we must avoid using those aggregate values to calculate a small numerical result. Thus, the more detailed results were used in this study.

Great awareness is suggested in viewing the results associated with missing values. Missing values do not imply zero or low. Specifically, when comparing by graphs, the missing values appear to be zero because they do not exist, or we may forget to pay attention to them. The missing value should be carefully interpreted again differently, such as by integrating several fields, reanalyzing them, or referring to other information.

Another important point is that mortality outcomes are not necessarily bad outcomes. Analyses that use postoperative death or survival as an outcome can be primarily those that equate with the “successfulness of surgery or perioperative” to some degree. For example, tracheostomies are often performed to continue the treatment of patients with respiratory disorders. Then, the result refers to whether death or survival can often be independent from surgical technique and perioperative management. This can be true for many other surgeries. For example, a patient who would otherwise have died if a certain surgery, e.g., coronary artery bypass surgery, was not performed would end up dying as a result of the procedure.

Nevertheless, comparisons of outcomes can and should be discussed from various perspectives. We can also consider whether a surgery should have been performed even if there were “no” other possible treatments. On the contrary, there may exist areas with inadequate medical care that cannot be implemented until the last resort. Thus, regarding outcomes in quality of care, background information of cases like case-mix must be considered. Such a detailed study interpretation requires a more detailed study with a more detailed protocol; however, it is very important to first present the current status, such as in this study, and to keep the interpretation flexible.

Other minor details include not a few inconsistencies in the data. For example, the K code may contain a 2-byte character “K” that is different from the original code, and data contained codes that do not exist. Thus, the conversion and unification of these characters can be considered when utilizing the dataset. Although “blood transfusion” is found in data that should not be reported as surgeries in Form 1 of DPC data by input rules, unlike pre-maintained databases such as registry databases, caution should be exercised in using unclean raw data. This study requested data acquisition according to the rules in place at the beginning of the study, and the handling of data inconsistencies and small numerical cells as described above had to be planned. Subsequently, it is now possible to conduct the study using individual data, which should allow for a more flexible analysis in the future.

## Conclusions

This study analyzed surgical outcomes at hospitals throughout Japan by region and over time using the highly comprehensive DPC data, provided comprehensive result data, and depicted regional differences and changes over time.

## Data Availability

All data produced in the present work are contained in the manuscript

## References

1. Uramoto H, Okumura M, Endo S, Tanaka F, Yokomise H, Masuda M. The 30-day mortality and hospital mortality after chest surgery described in the annual reports published by the Japanese Association for Thoracic and Cardiovascular Surgery. Gen Thorac Cardiovasc Surg. 2015;63:279–83.

2. Shiraishi Y. Current status of nontuberculous mycobacterial surgery in Japan: analysis of data from the annual survey by the Japanese Association for Thoracic Surgery. Gen Thorac Cardiovasc Surg. 2016;64:14–7.

3. Kumamaru H, Takahashi A, Fukuchi E, Ichihara N, Hirahara N, Miyata H. 2. National Clinical Database: Its Use and Data Quality Management Efforts. Japanese Journal of Pharmacoepidemiology/Yakuzai ekigaku [Internet]. 2016 [cited 2022 Jul 31];21:27–35. Available from: https://www.jstage.jst.go.jp/article/jjpe/21/1/21_27/_article/-char/ja/

4. Kakeji Y, Udagawa H, Unno M, Endo I, Kunisaki C, Taketomi A, et al. Annual Report of National Clinical Database in Gastroenterological Surgery 2015. The Japanese Journal of Gastroenterological Surgery. 2017;50:166–76.

5. Ando H, Yamaji K, Kohsaka S, Ishii H, Wada H, Yamada S, et al. Japanese Nationwide PCI (J-PCI) Registry Annual Report 2019: patient demographics and in-hospital outcomes. Cardiovasc Interv Ther. 2022;37:243–7.

6. Endo S, Ikeda N, Kondo T, Nakajima J, Kondo H, Yokoi K, et al. Development of an annually updated Japanese national clinical database for chest surgery in 2014. Gen Thorac Cardiovasc Surg. 2016;64:569–76.

7. Mizuno S, Kunisawa S, Sasaki N, Fushimi K, Imanaka Y. Effects of night-time and weekend admissions on in-hospital mortality in acute myocardial infarction patients in Japan. PLoS One. 2018;13:e0191460.

8. Bun S, Kunisawa S, Sasaki N, Fushimi K, Matsumoto K, Yamatani A, et al. Analysis of concordance with antiemetic guidelines in pediatric, adolescent, and young adult patients with cancer using a large-scale administrative database. Cancer Med. 2019;8:6243–9.

9. Okuno T, Kunisawa S, Fushimi K, Imanaka Y. Intra-operative autologous blood donation for cardiovascular surgeries in Japan: A retrospective cohort study. PLoS One. 2021;16:e0247282.

10. Watanabe S, Shin J, Morishita T, Takada D, Kunisawa S, Imanaka Y. Medium-Term Impact of the Coronavirus Disease 2019 Pandemic on the Practice of Percutaneous Coronary Interventions in Japan. J Atheroscler Thromb. 2021;63194.

11. Morishita T, Takada D, Shin J, Higuchi T, Kunisawa S, Fushimi K, et al. Effects of the COVID-19 pandemic on heart failure hospitalizations in Japan: interrupted time series analysis. ESC Heart Fail. 2022;9:31–8.

12. Kunisawa S, Imanaka Y. Regional Comparison of Quality of Care Using DPC Data (in Japanese). Shakaihokenjunpo [Internet]. 2019 [cited 2022 Aug 1];24–30. Available from: http://export.jamas.or.jp/dl.php?doc=5e4984c3339361af934a02f8ef6e2473379825c799c8273aa3a01163cbe9c0b9_bibtex.bib

13. Endo S, Ikeda N, Kondo T, Nakajima J, Kondo H, Yokoi K, et al. Model of lung cancer surgery risk derived from a Japanese nationwide web-based database of 78 594 patients during 2014-2015. Eur J Cardiothorac Surg [Internet]. 2017;52:1182–9. Available from: http://www.ncbi.nlm.nih.gov/pubmed/28977408

14. Kakeji Y, Takahashi A, Hasegawa H, Ueno H, Eguchi S, Endo I, et al. Surgical outcomes in gastroenterological surgery in Japan: Report of the National Clinical Database 2011-2018. Ann Gastroenterol Surg [Internet]. 2020;4:250–74. Available from: http://www.ncbi.nlm.nih.gov/pubmed/32490340

